# The influence of transtibial prosthesis type on gait adaptation: a case study

**DOI:** 10.1101/2022.06.27.22276778

**Authors:** Yosra Cherni, Simon Laurendeau, Maxime Robet, Katia Turcot

## Abstract

**Purpose:** Gait parameters are altered and asymmetrical in individual with transtibial amputation. The purpose of this study was to evaluate and compare the effect of four different prosthetic feet on lower-limb biomechanics during gait.

**Methods:** One young adult with transtibial ampution performed four gait analysis sessions with four foot-ankle prosthesis (Variflex, Meridium, Echelon, Kinterra). Kinematic, kinetic parameters and gait symmetry were analyzed during different prosthesis conditions.

**Results:** The type of prosthesis had little effect on amputee’ spatiotemporal parameters. Throughout the stance phase, an increase hip angle and a reduced knee flexion and ankle dorsiflexion were observed in the amputated leg. For kinetic parameters, a reduced propulsive force (SI=0.42–0.65), reduced knee extension moment (mainly during Echelon and Kinterra conditions, SI=0.17 and 0.32, respectively) and an increased knee abduction moment (mainly during the Variflex and Meridium, SI=5.74 and 8.93, respectively) in the amputated leg. Lower support moments were observed in the amputated leg compared to the unaffected leg, regardless of the type of prosthesis (SI=0.61–0.80).

**Conclusions:** The prostheses tested induced different lower-limb mechanical adaptations. If better gait symmetry between lower limbs is one of the clinical goals, an objective gait analysis could help clinicians to prescribe prosthetic feet based on quantitative measurement indicators.

## I. Introduction

Lower limb amputations can result from different etiology (*e.g*., traumatic, vascular, congenital, infection, *etc*). One of the most common amputations among lower limb is transtibial amputation (TTA) [1,2]. TTA occurs below the knee joint and deprives patients of the foot/ankle complex. The loss of the ankle and foot has a significant impact on the alteration of functional capacities and requires adaptation strategies to perform daily life activities, including walking and standing [3].

Compared to healthy individuals, individuals with TTA exhibit a large variety of functional limitations during gait due to the mechanical constraints imposed by their prosthesis [4,5]. These limitations are manifested mainly by a reduced walking speed [6], less generated power in the amputated leg during the push off phase [7], and increased inter-limb asymmetry in terms of spatiotemporal, kinematic and kinetic parameters (*e.g*., range of motion (ROM), external joint moments, ground reaction forces) [5]. Therefore, individuals with TTA must adapt their body-mechanical strategies to overcome these functional limitations and to ensure forward body progression during the swing phase. In addition, a risk of falling has also been recognised and associated with these functional limitations in individuals with TTA [8]. Because of these acquired gait body-mechanical strategies and asymmetries, individuals with TTA are more likely to develop secondary physical conditions (*e.g*., knee osteoarthritis, back pain, balance deficits) which can affect community integration and quality of life [9,10]. A group of stakeholders have identified five research action plans, including the importance to improve mobility which can result in decreasing functional limitations and promoting physical activity in this population [11].

Last decades, numerous ankle-foot prosthetic categories have been developed to address functional limitations in individuals with TTA. Despite their different mechanisms of action, the goal of these protheses is to restore the natural ankle–foot function to improve gait performances and reduce risk of tripping and falling [12]. For example, the majority of passive prosthetic ankle-foot components, such as energy storing and return prosthetic feet (*e.g*., Variflex), do not incorporate an articulating ankle joint [13]. Later, the ankle-foot component design has improved, with passive hydraulic ankle-foot prosthesis providing greater ROM and improved toe-clearance (*e.g*., Echelon and Kinterra) [12] and microprocessor-controlled prosthetic which automatically adapts to different types of walking and provides powered push-off (*e.g*., Meridium) [14]. However, the effect of these different types of prostheses on full lower-body gait adaptation has never been compared. The aim of this case study was then to evaluate the effect of four types of prostheses on body-mechanical strategies on the lower limbs in a participant with TTA (1) to identify which biomechanical variables are more influenced by the prosthesis type, and (2) to generate hypotheses with the goal to develop further studies. As the microprocessor-controlled prosthetic feet has been designed to increase ROM and to provide powered push-off in the amputated leg, it is expected that the Meridium would have a better influence on legs symmetry compared to other type of protheses. As gait rehabilitation process is a patient-specific treatment, a case study might be appropriate to better understand the effect of different prostheses.

## II. Materials and methods

### 1. Case description

A healthy and active man with unilateral TTA (age = in his 30s; weight = 81.8 kg; height = 1.7 m) participated to this study, within the framework of his medical supervision. His amputation was at the age of 2 months old due to a clogged artery and was made 6 cm below the knee cap. The participant was is able to walk without an assistive device. The participant provided written informed consent for participating in this study and for publishing the results.

### 2. Experimental protocol and data collection

The descriptions of different prostheses are reported in **Figure 1** and **Table S1**. To compare the effect of prostheses’ type on gait parameters, the participant performed four gait analysis sessions at a self-selected speed with four specific foot-ankle prosthesis (*i.e*., Variflex, Meridium, Echelon and Kinterra). All prostheses were checked for alignment and socket fit by an experienced prosthetist. For each prosthesis, a familiarization period of 7 to 14 days was completed.

**Figure 1.**
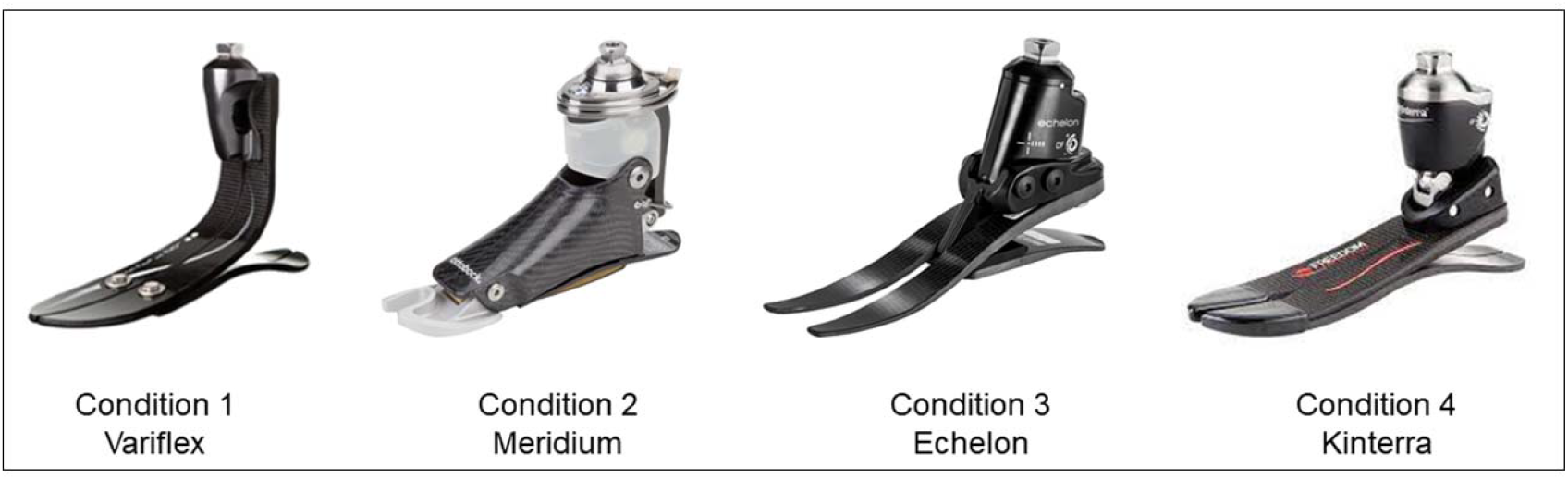
The four tested prostheses. Variflex LP (Ossur, Reykjavik, Iceland): ESR prosthesis with a carbon foot. Meridium (Ottobock, Duderstadt, Germany): a microprocessor-controlled prosthetic that automatically adjusts to different types of walking, changes in inclination and different walking speeds. Echelon VT (Chas. A Blatchford & Sons, Basingstoke, UK): A dynamic carbon fibre foot comprising independent toe and heel springs with a hydraulic self-aligning ankle. Kinterra (Freedom Innovations, Morgan, CA, USA): A hydraulic articulation combined with an energy-storage-and-return foot.

#### Gait analysis

A 10-cameras motion capture system (Vicon, Vantage 5, Oxford, UK) was used to collect the trajectories of reflective markers for kinematic data (sampled at 100 Hz) and four strain gauge force platforms embedded in the laboratory walkway (AMTI®, OR6, Watertown, MA, USA) were used for kinetic data (sampled at 1000 Hz). For the experimental calibration procedures, 72 reflective markers were placed bilaterally (and symmetrically on the prosthesis) using a 6 degrees of freedom model based on the International Society of Biomechanics recommendations for the lower limbs and the trunk [15]. The participant was asked to walk on a 10-m walkway at his self-selected comfortable speed wearing his own neutral shoes (same for each visit). Successful trials are those comprising a full contact on a single force platform and all makers viewable.

#### Prosthetic Profile of Amputee questionnaire

At the end of evaluation session, the Prosthetic Profile of Amputee (PPA) questionnaire was completed by the participant in order to identify the factors related to prosthesis use (*i.e*., comfort, adaptation, appearance, *etc*) [16]. Thus, the questionnaire was completed for each tested prosthesis. However, because the PPA is an extensive questionnaire covering various aspects which makes it lengthy, only items about the prosthetic were selected for this study.

### 3. Data analysis

#### Kinematic and kinetic parameters

Marker trajectories were tracked and analyzed using Nexus version 2.6.1 (Vicon Motion Systems, Oxford, UK). The MotionMonitor software (Innovative Sports Training, Chicago, IL, USA) was used to process spatiotemporal, kinematic, and kinetic data. Ankle, knee, hip and pelvis were calculated using a XYZ Cardan-Euler sequence. The pelvis segment angles were calculated with the respect to laboratory coordinate systems while the hip, knee and ankle angles and moments were calculated relative to the proximal segment. Inverse dynamic was used to calculate net external moments of hip, knee and ankle joints (all normalized by body mass). The vertical and anteroposterior ground reaction forces were measured by 2 piezoelectric force plates, placed in the middle of the 10-m walkway of the laboratory. For each prosthesis condition, 8 to 16 gait cycles were selected for analysis using a customized software application (Moveck Solution Inc, Quebec, Canada).

#### Total Support Moment

As defined by Winter [17], the total support moment (TSM) was calculated as the sum of extensor moments of the ankle, knee, and hip at the entire stance phase as well as the subphases of stance.

#### Gait symmetry

The symmetry index (SI) compared the kinematics and kinetics of the amputated leg to the unaffected leg for each type of prosthesis tested. The symmetry index was calculated during the stance phase of the gait cycle for each subject and was calculated for each condition, as follows [18,19]:

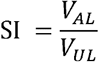

with *V*_*AL*_ and *V*_*UL*_ represent the values of the gait parameters, calculated, respectively, for the amputated leg and the unaffected leg. When no differences are measured between the two limbs, SR equals one, while lower or higher values indicates asymmetry in the gait parameters. Using a ratio equation to describe asymmetry is recommended when there is an identifiable affected side, as the results are relatively easy to interpret [20,21].

## III. Results

### 3.1. Prosthetic Profile of the Amputee

The results of prosthesis use section of PPA are displayed in **Table S1**. Overall, the participant was more satisfied with the use of the Echelon in terms of comfort, weight, appearance, and gait appearance compared to other prostheses. In terms of comfort, the Meridium was less comfortable than other prostheses (**Table S1**).

### 3.2. Kinematic parameters

<u>Spatiotemporal parameters</u> are summarized in **Table 1**. Overall, the tested prostheses allowed a symmetry of the spatiotemporal parameters ranging from 0.94 to 1.03, except for the step length when walking with Kinterra prosthesis (SI = 1.25).

**Table 1.** Kinematics and kinetics’ outcomes during the four conditions. Values are expressed as mean (SD).

|  | Amputated leg |  |  |  | Unaffected leg |  |  |  |
| --- | --- | --- | --- | --- | --- | --- | --- | --- |
|  | Variflex | Meridium | Echelon | Kinterra | Variflex | Meridium | Echelon | Kinterra |
| **Spatiotemporal parameters** |  |  |  |  |  |  |  |  |
| Speed (m/s) | 1.20 (0.03) | 1.15 (0.03) | 1.13 (0.02) | 1.15 (0.03) | 1.19 (0.02) | 1.16 (0.02) | 1.13 (0.02) | 1.17 (0.03) |
| Step length (m) | 0.67 (0.03) | 0.69 (0.02) | 0.67 (0.01) | 0.69 (0.02) | 0.67 (0.03) | 0.70 (0.01) | 0.67 (0.01) | 0.55 (0.28) |
| Cadence (step/min) | 105.30 (2.36) | 100.02 (1.75) | 101.13 (0.90) | 101.05 (0.42) | 105.16 (1.34) | 100.85 (1.20) | 101.41 (1.03) | 101.94 (2.15) |
| Stance (%) | 62 (0.82) | 61 (1.03) | 63 (0.30) | 65 (4.02) | 64 (0.71) | 65 (0.93) | 66 (0.37) | 63 (0.75) |
| **Joint angles** |  |  |  |  |  |  |  |  |
| Pelvis Ante/Retroversion (°) |  |  |  |  |  |  |  |  |
| - ROM at stance | 7.67 (1.11) | 9.56 (1.47) | 8.92 (0.54) | 8.51 (3.02) | 9.17 (1.10) | 8.98 (1.29) | 8.74 (1.01) | 10.28 (1.37) |
| - Peak anteversion at stance | 6.40 (1.20) | 5.23 (0.67) | 8.39 (0.60) | 2.18 (1.92) | 7.62 (0.85) | 5.21 (0.85) | 9.08 (0.53) | 3.47 (1.43) |
| - Peak retroversion at stance | -1.26 (0.46) | -4.32 (0.95) | -0.53 (0.72) | -6.33 (1.30) | -1.55 (0.72) | -3.78 (0.57) | 0.34 (0.39) | -6.81 (0.99) |
| Hip Flexion/Extension (°) |  |  |  |  |  |  |  |  |
| - ROM at stance | 44.66 (2.13) | 59.08 (2.20) | 53.04 (1.15) | 47.74 (7.57) | 42.37 (1.01) | 38.48 (1.09) | 37.65 (1.32) | 41.83 (1.11) |
| - Peak flexion at stance | 25.18 (1.09) | 33.25 (1.95) | 30.77 (1.27) | 26.62 (4.65) | 22.35 (1.08) | 27.88 (1.21) | 22.80 (1.18) | 33.59 (1.05) |
| - Peak extension at stance | -19.47 (1.32) | -25.83 (1.42) | -22.26 (1.19) | -21.12 (4.70) | -20.02 (0.40) | -10.60 (1.01) | -14.85 (0.67) | -8.24 (1.12) |
| Knee Flexion/Extension (°) |  |  |  |  |  |  |  |  |
| - ROM at stance | 55.37 (2.23) | 37.46 (2.53) | 44.17 (0.94) | 59.24 (4.45) | 35.74 (3.5) | 33.78 (2.48) | 33.92 (3.50) | 28.33 (3.31) |
| - Peak flexion at stance | 51.24 (2.30) | 30.48 (2.50) | 40.15 (0.93) | 47.74 (3.72) | 31.27 (3.98) | 33.34 (2.36) | 35.58 (2.15) | 36.23 (2.94) |
| - Peak extension at stance | -4.13 (0.27) | -6.98 (0.23) | -4.02 (0.17) | -11.49 (1.32) | -4.46 (1.70) | -0.45 (1.24) | 1.66 (1.64) | 7.90 (1.17) |
| Ankle Flexion/Extension (°) |  |  |  |  |  |  |  |  |
| - ROM at stance | 22.92 (0.53) | 18.23 (0.81) | 14.82 (0.37) | 22.02 (0.92) | 21.87 (2.26) | 25.23 (1.35) | 24.63 (1.14) | 25.06 (1.26) |
| - Peak dorsiflexion at stance | 19.92 (0.36) | 16.08 (1.04) | 13.35 (0.36) | 20.37 (1.29) | 7.05 (0.54) | 13.19 (0.68) | 9.27 (0.75) | 11.08 (1.63) |
| - Peak plantar flexion at stance | -3.00 (0.22) | -2.15 (1.25) | -1.47 (0.26) | -1.65 (1.61) | -14.83 (2.33) | -12.04 (1.33) | -15.36 (1.28) | -13.98 (0.38) |
| **Kinetic parameters** |  |  |  |  |  |  |  |  |
| Ground reaction force (N/kg) |  |  |  |  |  |  |  |  |
| - 1<sup>st</sup> vertical peak | 10.01 (0.61) | 9.88 (0.29) | 9.79 (0.20) | 10.65 (0.46) | 10.73 (0.09) | 10.40 (0.22) | 9.84 (0.18) | 10.47 (0.21) |
| - 2<sup>nd</sup> vertical peak | 10.40 (0.07) | 11.11 (0.13) | 10.41 (0.35) | 10.73 (0.12) | 9.50 (0.07) | 9.70 (0.21) | 9.52 (0.06) | 9.04 (0.11) |
| - AP Braking peak | -1.58 (0.06) | -1.41 (0.23) | -1.47 (0.08) | -1.92 (0.12) | -1.86 (0.23) | -1.27 (0.05) | -1.63 (0.01) | -1.49 (0.17) |
| - AP Propulsion peak | 1.28 (0.07) | 0.88 (0.06) | 1.19 (0.15) | 1.22 (0.05) | 1.96 (0.08) | 2.10 (0.02) | 2.00 (0.05) | 2.01 (0.03) |
| Joint moments (Nm/kg) |  |  |  |  |  |  |  |  |
| - Peak hip extension | -0.46 (0.01) | -0.63 (0.06) | -0.53 (0.08) | -0.53 (0.03) | -0.75 (0.10) | -1.02 (0.02) | -0.81 (0.07) | -0.79 (0.02) |
| - Peak knee extension | -0.30 (0.02) | -0.43 (0.03) | -0.15 (0.02) | -0.07 (0.01) | -0.46 (0.03) | -0.43 (0.01) | -0.46 (0.06) | -0.42 (0.03) |
| - Peak ankle plantarflexion | 0.86 (0.02) | 1.16 (0.01) | 0.98 (0.06) | 1.08 (0.02) | 1.03 (0.01) | 1.02 (0.01) | 1.13 (0.01) | 1.10 (0.01) |
| - Peak hip abduction at early stance | 0.74 (0.08) | 0.82 (0.07) | 0.68 (0.03) | 0.88 (0.04) | 0.67 (0.07) | 0.68 (0.01) | 0.66 (0.05) | 0.77 (0.02) |
| - Peak hip abduction at late stance | 0.87 (0.06) | 0.94 (0.09) | 0.62 (0.05) | 0.84 (0.02) | 0.48 (0.01) | 0.50 (0.07) | 0.63 (0.01) | 0.58 (0.02) |
| - Peak knee abduction at early stance | 0.46 (0.06) | 0.56 (0.03) | 0.40 (0.03) | 0.57 (0.04) | 0.22 (0.02) | 0.20 (0.01) | 0.41 (0.03) | 0.25 (0.05) |
| - Peak knee abduction at late stance | 0.46 (0.05) | 0.49 (0.03) | 0.32 (0.01) | 0.40 (0.02) | 0.08 (0.01) | 0.05 (0.03) | 0.26 (0.02) | 0.13 (0.01) |
| Total support moment (Nm/kg) |  |  |  |  |  |  |  |  |
| - TMS | 1.97 (0.01) | 2.40 (0.23) | 2.24 (0.02) | 2.17 (0.10) | 2.77 (0.23) | 2.99 (0.17) | 2.86 (0.11) | 3.53 (0.14) |
| - Hip contribution | 0.53 (0.01) | 0.67 (0.15) | 0.66 (0.04) | 0.67 (0.10) | 0.55 (0.06) | 0.54 (0.02) | 0.41 (0.03) | 0.45 (0.02) |
| - Knee contribution | 0.25 (0.02) | 0.29 (0.05) | 0.09 (0.01) | 0.05 (0.01) | 0.43 (0.05) | 0.53 (0.05) | 0.68 (0.03) | 0.70 (0.12) |
| - Ankle contribution | 1.20 (0.01) | 1.44 (0.05) | 1.49 (0.04) | 1.46 (0.05) | 1.80 (0.18) | 1.92 (0.21) | 1.77 (0.07) | 2.38 (0.16) |

<u>Joint angles results</u> are displayed in **Table 1** and **Figure 2**. In <u>the sagittal plane</u>, an increase in pelvic retroversion was observed during the stance phase when walking with the Kinterra prosthesis provoking a greater hip flexion in the unaffected leg compared to other conditions. Knee extension was observed in the amputated leg throughout all the stance phase during the four prostheses conditions, but more prominent with the Kinterra prosthesis (knee extension = 11.49°). During all prosthesis’s conditions, increased ankle dorsiflexion for the amputated leg with an absence of push-off on were observed compared to the unaffected leg. In terms of gait symmetry, walking with with the Variflex and Kinterra prostheses induced lower ROM symmetry at the pelvis (SI = 0.84 and 0.82, respectively) and knee (SI = 1.55 and 2.09, respectively) during the stance phase. The Meridium and Echelon prostheses tend to increase the hip ROM of the amputee leg during the stance phase (SI = 1.51 and 1.41, respectively). In <u>the frontal plane</u> (**Figure 2**), a greater pelvis elevation at the amputated side was observed when walking with the Kinterra prosthesis. Furthermore, with the latter prosthesis, the prosthetic ankle maintained an adducted position throughout the gait cycle.

**Figure 2.**
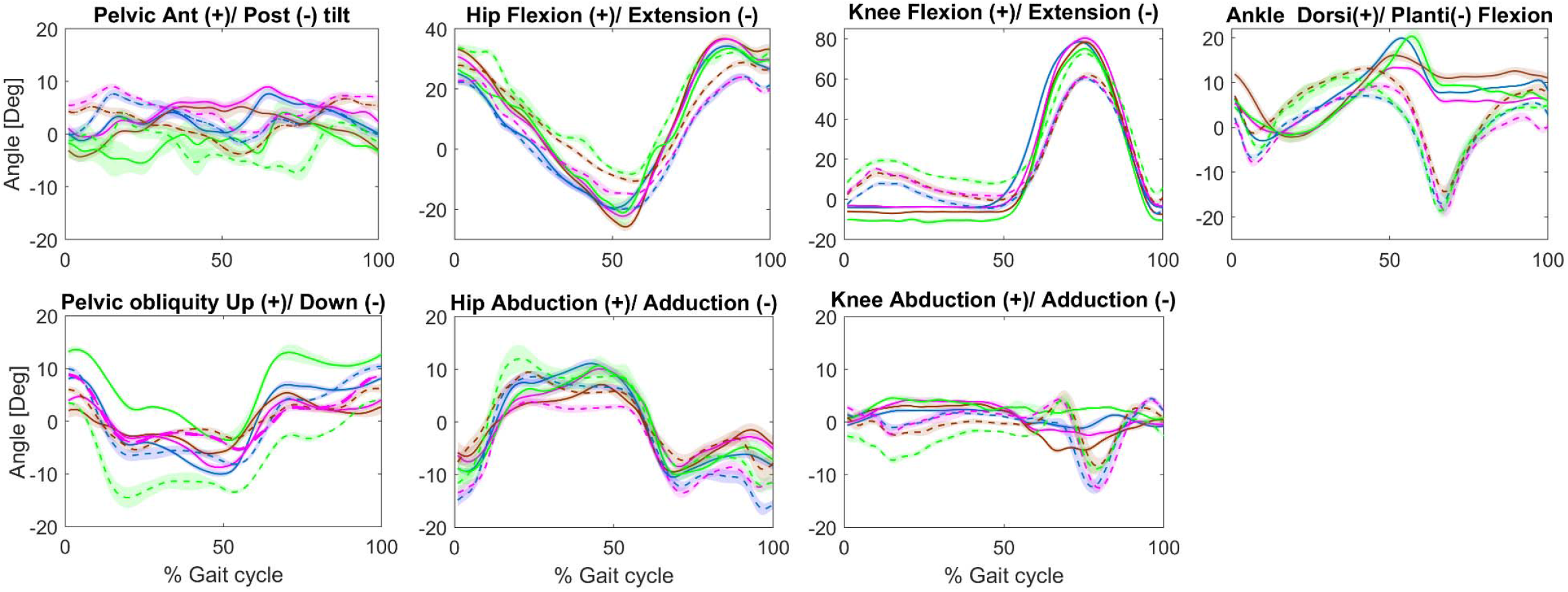
Joint angles in the sagittal and frontal plane during walking in four prostheses conditions: Variflex (blue), Meridium (brown), Echelon (magenta) and Kinterra (green). Kinematic of amputated leg presented in continuous line and that of the unaffected leg presented in dotted line. Shaded region denotes standard deviation.

### 3.3. Kinetic parameters

<u>The results of the ground reaction force</u> from the amputated and unaffected legs during different prostheses’ conditions are shown in **Table 2** and **Figure 3**. Regarding the vertical component of the ground reaction force, differences between the legs reflected mainly higher peak force in late stance the amputated leg compared to unaffected leg (SI = 1,09 – 1.19). This difference in the second vertical peak force was especially apparent at the Meridium and Kinterra conditions (SI = 1.15 and 1.19, respectively). For the anterior–posterior component of the ground reaction force, the amputated leg had lower peak propulsive forces than the unaffected leg (SI = 0.42 – 0.65) during all the prostheses’ conditions. The difference in peak of propulsive forces between the amputated and unaffected legs was greater during the Meridium condition (SI = 0.42).

**Table 2.** Symmetry index for kinematic and kinetic outcomes during the four conditions.

|  | SI-Variflex | SI-Meridium | SI-Echelon | SI-Kinterra |
| --- | --- | --- | --- | --- |
| **Spatiotemporal parameters** |  |  |  |  |
| Speed | 1.01 | 0.99 | 1.00 | 0.98 |
| Step length | 1.00 | 0.99 | 1.00 | 1.25 |
| Cadence | 1.00 | 0.99 | 1.00 | 0.99 |
| Stance phase | 0.96 | 0.94 | 0.95 | 1.03 |
| **Sagittal joint angles** |  |  |  |  |
| - Pelvis ROM at stance | 0.84 | 1.06 | 1.02 | 0.82 |
| - Hip ROM at stance | 1.05 | 1.54 | 1.41 | 1.14 |
| - Knee ROM at stance | 1.55 | 1.11 | 1.30 | 2.09 |
| - Ankle ROM at stance | 1.05 | 0.72 | 0.60 | 0.88 |
| **Kinetic parameters** |  |  |  |  |
| Ground reaction force |  |  |  |  |
| - 1<sup>st</sup> vertical peak | 0.93 | 0.95 | 0.99 | 1.02 |
| - 2<sup>nd</sup> vertical peak | 1.09 | 1.15 | 1.09 | 1.19 |
| - AP Braking peak | 0.85 | 1.11 | 0.90 | 1.29 |
| - AP Propulsion peak | 0.65 | 0.42 | 0.60 | 0.61 |
| Joint moments (Nm/kg) |  |  |  |  |
| - Peak hip extension | 0.61 | 0.62 | 0.65 | 0.66 |
| - Peak knee extension | 0.66 | 0.99 | 0.32 | 0.17 |
| - Peak ankle plantiflexion | 0.83 | 1.14 | 0.89 | 0.98 |
| - Peak hip abduction at early stance | 1.09 | 1.21 | 1.03 | 1.15 |
| - Peak hip abduction at late stance | 1.80 | 1.88 | 0.99 | 1.45 |
| - Peak knee abduction at early stance | 2.14 | 2.81 | 0.98 | 2.32 |
| - Peak knee abduction at late stance | 5.71 | 8.93 | 1.22 | 3.17 |
| Total support moment |  |  |  |  |
| - TMS in stance | 0.71 | 0.80 | 0.79 | 0.61 |
| - Hip contribution to TMS | 0.96 | 1.25 | 1.61 | 1.47 |
| - Knee contribution to TMS | 0.57 | 0.55 | 0.13 | 0.07 |
| - Ankle contribution to TMS | 0.67 | 0.75 | 0.84 | 0.61 |

**NOTE:** ROM: Range of Motion; AP: Antero-posterior; TSM: Total Moment Support; SI: Symmetry Index. with a value of 1 indicating perfect. SI > 1 indicate asymmetry with higher parameter value for the amputated leg, and values < 1 indicate asymmetry with higher parameter value for the unaffected leg.

**Figure 3:**
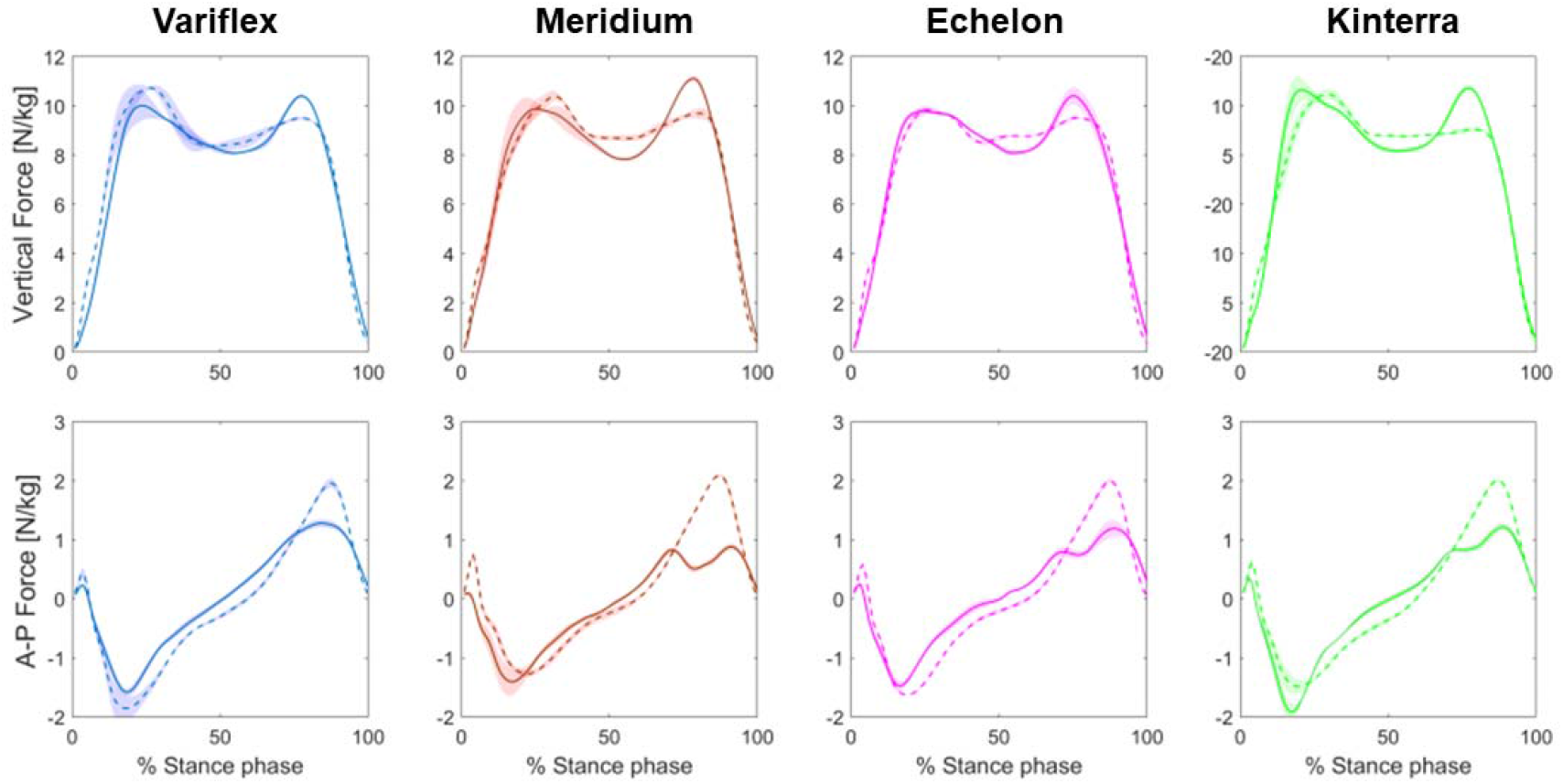
Mean vertical and anterior–posterior components of the ground reaction force patterns during stance at different prostheses’ conditions. Forces of amputated leg presented in continuous line and that of the unaffected leg presented in dotted line. Shaded region denotes standard deviation.

<u>Regarding joint moments</u>, the hip extension moment decreased at late stance in the amputated leg in all the tested conditions (**Figure 4**). Relative to the unaffected leg, a lack of knee extension moment was observed in the amputated leg during stance. Compared to the unaffected leg, ankle plantarflexion moment was decreased during stance in the amputated leg when walking with the Variflex and Echelon prostheses (SI = 0.83 – 0.89). In regards of the frontal plane, all prothesis with the exception of the Echelon had increased the knee abduction moment in the amputated leg at early (SI = 2.14 – 2.81) and late stance (SI = 3.17 – 8.93). However, the Echelon prosthesis allowed a between limbs’ asymmetry at early (SI = 0.98) and late stance (SI = 1.22).

**Figure 4:**
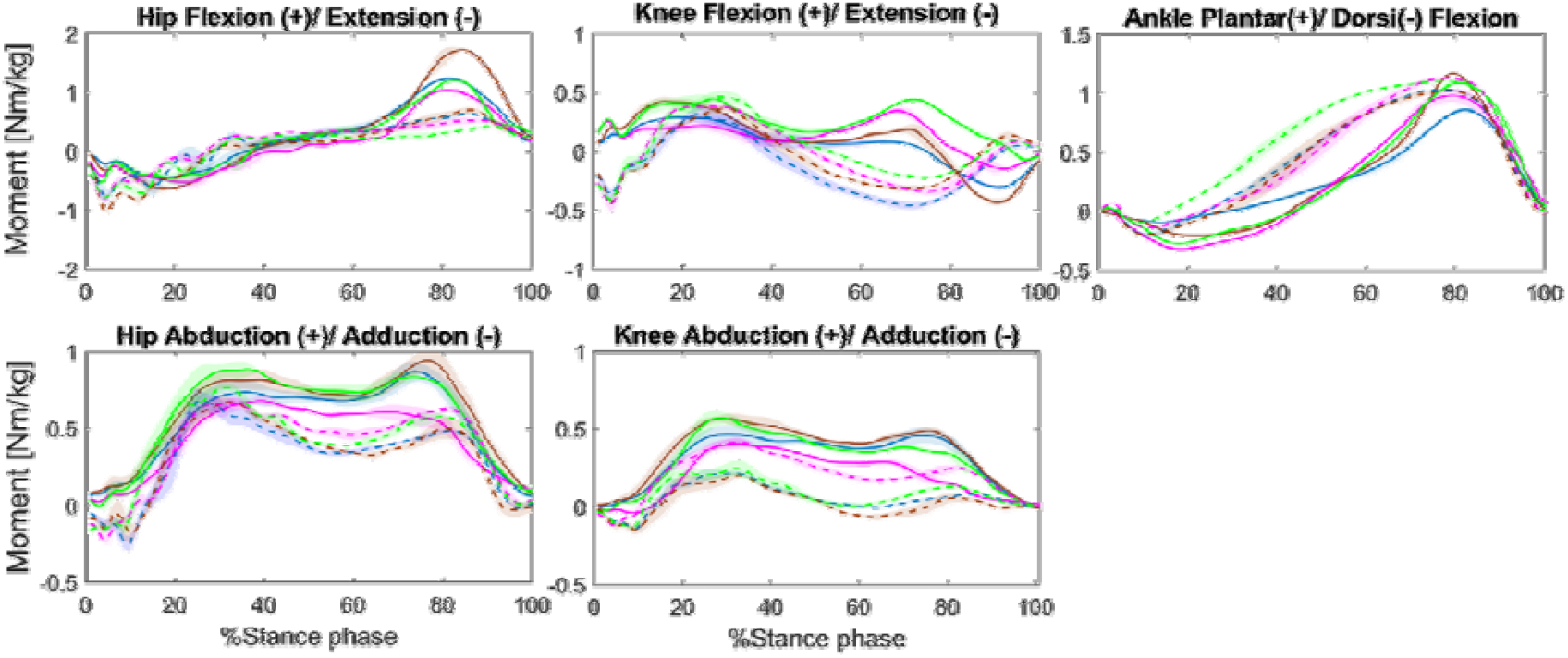
Joint moment mean waveforms for four prostheses’ conditions subject groups: Variflex (blue), Meridium (brown), Echelon (magenta) and Kinterra (green). Kinematic of amputated leg presented in continuous line and that of the unaffected leg presented in dotted line. Shaded region denotes standard deviation.

The combined effect of the net extensor moments of the hip, knee, and ankle joints are reflected by <u>the TSM</u> as showed in **Table 1** and **Figure 5**. Overall, the participant generated lower support moments in the amputated leg compared to the unaffected leg, regardless of the type of prosthesis (SI = 0.61 – 0.80). The TSM of the amputated leg was lower with the Variflex prosthesis compared to other prostheses. In terms of joint contributions to TSM, more substantial differences between limbs were apparent at the knee (**Figure 5)**, particularly during the Echelon (contribution = 4% in amputated leg ***vs*** 24% in unaffected leg) and Kinterra conditions (contribution = 2% in amputated leg ***vs*** 20% in unaffected leg).

**Figure 5:**
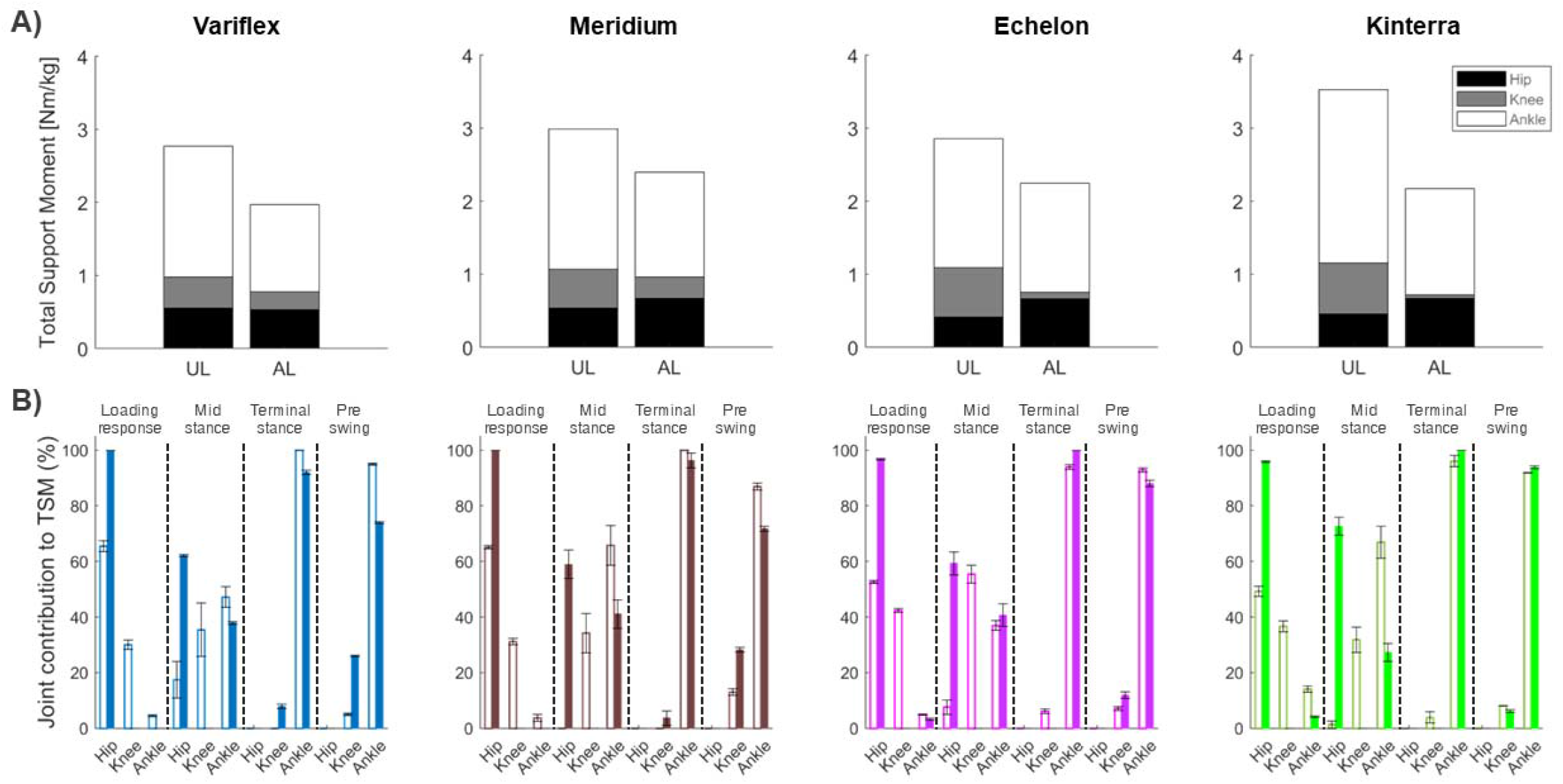
**(A)** Mean total support moment at the four conditions during the entire stance phase. **(B)** Individual joint contributions to total support moment as a function of the sub-phases of the stance phase. Results of amputated leg presented in filled bars and that of the unaffected leg presented in bars with colored borders: Variflex (blue), Meridium (brown), Echelon (magenta) and Kinterra (green).

## IV. Discussion

The objective of this case study was to compare the effect of four types of prostheses on biomechanical gait parameters in one participant with a TTA. A single-subject design was used in this study to preserve the participant-specific information as previously described in other studies [22,23]. Overall, the results of this study showed a disparity in biomechanical adaptations between the different prostheses. However, three major findings warrant to be noticed. Firstly, throughout the stance phase, an increase hip ROM and a reduced knee flexion and ankle dorsiflexion were observed in the amputated leg when walking, which highlights specific asymmetries induced by each prosthesis. Secondly, a reduced propulsive force (IS = 0.42 – 0.65), reduced knee extension moment (mainly during Echelon and Kinterra conditions, IS = 0.17 and 0.32, respectively) and an increased knee abduction moment (mainly during Variflex and Meridium conditions, IS = 5.71 and 8.93, respectively) were denoted in the amputated leg. Thirdly, the amputee generated lower support moments in the amputated leg compared to the unaffected leg, regardless of the type of prosthesis.

### Effect of different prostheses on lower limb kinematics

The Echelon and Meridium prostheses induced a greater hip ROM at the amputated side compared to the unaffected leg (SI ^=^ 1.41 – 1.54), causing a higher asymmetry. This result is probably the evidence of a compensation of the reduced ROM at the ankle when walking with these same prostheses. On the other hand, important knee ROM asymmetries were observed mainly during the Variflex and Kinterra conditions (SI = 1.55 – 2.09, respectively). Indeed, the residual knee remained in extension throughout the stance phase in the four prosthetic conditions, but more prominent during the Kinterra condition (knee extension = 11.49°). This could be due to insufficient socket flexion (foot plantar flexed) [24]. In addition, the loss of knee flexion in the amputated leg may require body-mechanical compensations which could affect load bearing in the intact knee. For example, the observed reduction in residual knee flexion angle during midstance could be evidence of a quadriceps-avoidance gait. Previous studies [25,26] reported a compensatory muscle activity in individuals with TTA during walking, namely asymmetry in intact *vs*. residual knee flexor/extensor activity. In general, abnormal knee kinematics are linked to the onset of knee osteoarthritis [27]. Similarly, a notable difference between legs occurred at the ankle joint, particularly in late stance and early swing when the plantar flexion movement within the prosthesis was substantially lower than that of the intact ankle joint, regardless of the prosthesis conditions. Increased ankle dorsiflexion of the amputated leg is a strategy that facilitates minimal toe clearance which is required to prevent tripping and falls. However, the lack of ankle plantarflexion as observed in all the conditions affects the propulsion. These kinematic deviations are coherent with the observed differences between legs in terms of joint moments and ground reaction forces.

### Effect of different prostheses on lower limb kinetics

In terms of joint moments, large asymmetries were present in maximum hip and knee extensor moments during gait suggesting that muscle coordination and braking/propulsion effort may be altered for the residual leg with different prostheses. Moreover, the knee adduction moment is an important parameter, as the peak knee adduction moment has been related to the initiation and progression of knee OA [28]. In the present study, the amputee showed a higher knee abduction moment in the residual knee when walking with the Meridium and Kinterra prostheses compared to other conditions, which leads the intact knee to a moment tending towards adduction. However, the Echelon tends to induce similar abduction moment at the intact and residual knee. For the vertical ground reaction forces, all conditions reflected the standard pattern with the double peaks. Our results showed that the magnitude of the 1^st^ peak was quite similar on both sides, whatever the prosthesis used. This was not the case for the 2^nd^ peak that increased in the amputated leg compared to the unaffected leg (mainly when walking with the Meridium or Kinterra). One possible explanation could be that the amputee prefers to load his weight on the unaffected leg, as a protective gait strategy. For the AP ground reaction forces, the propulsive peak was lower by 34% to 58% in the amputated leg (mainly when walking with the Meridium prosthesis). This result agrees with kinematics results that showed a more vertical leg orientation (as showed by an increase in hip and knee extension in the amputated leg) which would result in a reduction in the AP ground reaction forces, in addition to the inability of the amputee to modulate the ankle plantar flexor moment during stance (**Figure 4**). These deviations together contribute to a less propulsive gait [29].

The combined effect of the net extensor moments of the hip, knee, and ankle joints are reflected by the TSM. In general, a smaller TSM was observed for the amputated leg compared to the unaffected leg. This result may underline the inability of amputee to overcome the loss of ankle extensor muscles (*i.e*., Soleus and Gastrocnemus) that contribute to vertical support throughout single stance [30]. During mid stance phase, Soleus and Gastrocnemus together ensures support and forward progression of both the leg and trunk [30,31]. Thus, the loss or impairment of Soleus and Gastrocnemus force generation would impact gait stability. In addition, the results of this study showed a small contribution of the knee extensor moment to TSM in comparison with the ankle and hip joints in both legs. These results are consistent with previous studies that showed a dominant contribution of the ankle moment in support and propulsion during locomotion [17,32]. However, this effect was amplified when walking with Echelon or Kinterra prostheses for which the knee moment of the amputated leg never became extensors for all of the stance phase. Thus, a net knee extensor moment was not crucial to the development of the necessary extensor support moment on the amputated leg when walking with these prostheses. Similarly, Sanderson et al. [33] showed that TTA amputees were able to generate an extensor support moment on the amputated leg in the absence of the knee extensor moment. As showed by our results, the decrease in knee contribution seems to be mainly compensated for by the hip extensor moment. In summary, the results of this case suggest that depending on the type of prosthesis, the amputee adopted different support strategies in the unaffected and amputated legs.

### Clinical implications

In summary, locomotor adaptations were different from one prosthesis to another. However, the lesser need for proximal compensations at the hip and knee may be key factors underlying the preference. The present results demonstrated a better symmetry of lower limb joint moments (except for knee extensor moment) and ground reaction forces when using the Echelon prosthesis. However, specific to this case, the Kinterra condition resulted in higher pelvis and knee ROM asymmetries, which may reveal important proximal compensations in the unaffected leg. This result may be explained by the fact that this prosthesis offers less control of the prosthetic ankle and therefore induces more proximal compensations are needed. Similarly, large asymmetries were present in lower limb extension moments during gait mainly when walking with Echelon and Kinterra prostheses, suggesting that muscle coordination and braking/propulsion effort may be more affected in the amputated leg with these prostheses. Compared to unaffected leg, the Variflex and Meridium prostheses induced an important increase in residual knee abduction moments that could present an important risk factor for of knee OA. Finally, the patient preferences, needs and comfort as well as the prosthesis cost are also factors to consider. For example, the Meridium with the highest cost among the tested prostheses seems to be the least comfortable prosthesis for the participant (**Table S2**). However, given the participant profile (low displacement with the prosthesis) and specificity of this prosthesis (a microprocessor-controlled prosthesis, **Table S1**), a longer familiarization period would be necessary to overcome this discomfort. Finally, within the framework of this study, the choice was made on the Echelon which seems to answer better the need for the participant from a point of view comfort, gait adaptation, and cost.

### Study limitations

There are some limitations in this study that need to be acknowledged. Firstly, only one case was reported which reduces the external validity of the results. However, this design was used to generate hypotheses for further studies to verify if these results would translate to a larger population. Secondly, the present results are also limited to only walking task. Further studies assessing effect of different type prostheses during various functional tasks (*e.g*., sit to stand, stair ascent and descent, physical activity) are needed to better understand their functional contribution in a real-life situation. Thirdly, muscle activity during gait were not recorded which limits the understanding of the compensatory strategies induced by the different prostheses.

## V. Conclusion

Gait impairments in individuals with TTA can be attributed to either mechanical constraints imposed by the prosthesis or amputee-specific factors. The results of this case study showed a disparity in biomechanical adaptations between the different prostheses in terms of joint angles, moments, and contributions. Finally, objective and multifactorial determinants (*e.g*., environmental, contextual, workspace, *etc*) may be needed for prescribing prosthetic feet with respect to the functional contribution to amputee.

## Data Availability

All data produced in the present study are available upon reasonable request to the authors

## Supplementary materials

**Table S1.** Description of tested protheses.

|  | **Variflex LP**<br>**(Ossur)** | **Meridium**<br>**(Ottobock)** | **Echelon VT**<br>**(Blatchford)** | **Kinterra**<br>**(Freedom Innovations)** |
| --- | --- | --- | --- | --- |
| **Category** | Energy Storage and return foot | A microprocessor-controlled prosthetic | A dynamic carbon fibre foot comprising independent toe and heel springs with a hydraulic self-aligning ankle. | Combine a hydraulic articulation unit with an energy-storage-and-return foot |
| **Degrees of freedom** | The Variflex LP does not offer assisted ankle plantar/dorsal flexion. The patient must exert a force on the carbon plate which will induce an energy return. | A 4-axis mechatronic foot with hydraulic movement of the ankle. It offers 36.5 degrees of plantar-flexion and dorsal-flexion. | A hydraulic foot that offers 9 degrees of plantar-flexion and dorsi-flexion. | A hydraulic foot that offers 12 degrees of ankle mobility. |
| **Prosthesis specification** | This prosthesis was combined to DeltaTwist shock absorber, which also features a torsion function. | The Meridium automatically adjusts to different types of walking, changes in inclination and different walking speeds. | The Echelon VT offers shock absorption and a rotation adapter included in the foot. | The Kinterra foot was combined with a durashock. This piece of elastomer serves as a shock absorber and rotation adapter. |

**Table S2.** The prosthesis section results of the PPA questionnaire.

| Prosthesis section | Subsection | Variflex | Meridium | Echelon | Kinterra |
| --- | --- | --- | --- | --- | --- |
| **Question 1** | Comfort | 4 | 5 | 5 | 5 |
|  | Appearance | 5 | 5 | 5 | 5 |
|  | Weight | 5 | 3 | 5 | 5 |
|  | Appearance of your gait | 4 | 5 | 5 | 4 |
| **Question 2** | Amputation | 5 | 5 | 5 | 5 |
|  | Prosthesis | 5 | 3 | 5 | 4 |
| **Question 3** |  | None | Insecurity walking in a small environment due the inability of the leg to react quickly. | None | Insecurity when making turns or picking up objects if I must rotate. |
| **Question 4** |  | Changed for a different type of prosthesis | Changed for a different type of prosthesis | Changed for a different type of prosthesis | Changed for a different type of prosthesis |
| **Question 5** | Is able to quickly give you an appointment? | Yes | Yes | Yes | Yes |
|  | Is located sufficiently close to your home? | Yes | Yes | Yes | Yes |

**NOTE: Question 1:** Four characteristics concerning your prosthesis are listed below. Please indicate your degree of satisfaction for each one of these characteristics.; **Question 2:** The adaptation (in the sense of «GETTING used to...») to the amputation and to the prosthesis may be more difficult for some people than for others, and this adaptation is not always easy to evaluate. After examining the given choices of possible answers, please indicate the answer that best describes your actual adaptation to your; **Question 3:** At the present time, when you wear your prosthesis, does it cause you; **Question 4:** Since the time you completed your rehabilitation program, has your prosthesis been; **Question 5:** In your opinion, your prosthesis laboratory.

